# SARS-CoV-2 infection in fully vaccinated individuals of old age strongly boosters the humoral immune response

**DOI:** 10.1101/2021.07.19.21260563

**Authors:** Lisa Müller, Marcel Andrée, Philipp Niklas Ostermann, Nathalie Jazmati, Greta Flüh, Johannes Fischer, Edwin Bölke, Hilmar Wisplinghoff, Heiner Schaal, Ingo Drexler, Andreas Walker, Ortwin Adams, Jörg Timm

**Author notes:** Corresponding author: Lisa Müller, Corresponding author. contributed equally (Lisa Müller, Marcel Andrée, Philipp Niklas Ostermann).

## Abstract

**Introduction:** Prophylactic vaccination against SARS-CoV-2 is one of the most important measures to contain the COVID-19 pandemic. Recently, break-through infections following vaccination against this virus have been reported. Here, we describe the humoral immune response of break-through infections in fully vaccinated individuals of old age from an outbreak in a nursery home.

**Methods:** In cooperation with the local health authority, blood samples from fully vaccinated and infected as well as fully vaccinated and uninfected residents of the nursery home were collected four weeks after the onset of the outbreak. The humoral immune response was determined in a neutralisation assay with replication-competent virus isolates and by a quantitative ELISA.

**Results:** In this outbreak a total of 23 residents and four health care workers were tested positive for SARS-CoV-2. Four residents were unvaccinated, including one with a severe course of disease who later deceased. Despite their old age, all vaccinated residents showed no or only mild disease. Comparison of the humoral immune response revealed significantly higher antibody levels in fully vaccinated infected individuals compared to fully vaccinated uninfected individuals (p<0.001). Notably, although only a minority of the vaccinated uninfected group showed neutralisation capacity against SARS-CoV-2, all vaccinated and infected individuals showed high-titer neutralisation of SARS-CoV-2 including the alpha and beta variant.

**Discussion:** Large SARS-CoV-2 outbreaks can occur in fully vaccinated populations, but seem to associate with mild disease. SARS-CoV-2 infection in fully vaccinated individuals is a strong booster of the humoral immune response providing enhanced neutralisation capacity against immune evasion variants.

## Introduction

Prophylactic vaccination against SARS-CoV-2 is one of the most important measures to contain the COVID-19 pandemic (Bagchi, Mak et al. 2021, Britton, Jacobs Slifka et al. 2021, Dagan, Barda et al. 2021, Gharpure, Guo et al. 2021, Thompson, Burgess et al. 2021). The vector vaccines and mRNA-based vaccines approved in Europe have been able to show a protective effect against symptomatic infections in the respective phase 3 studies (Zhu, Zhang et al. 2020, Dagan, Barda et al. 2021). Effectiveness studies also documented a very good protective effect against hospitalisation as an indicator of a severe course of infection as early as four weeks after the first vaccine dose (Hall, Foulkes et al. 2021). In cases where an infection occurred after vaccination, the concentration of viral RNA in swabs was significantly lower and the secondary attack rate from vaccinated index persons was substantially reduced compared to unvaccinated index persons (Haas, Angulo et al. 2021, Levine-Tiefenbrun, Yelin et al. 2021). Thus, in addition to individual protection, vaccination can also contribute to protection against the spread of SARS-CoV-2. In Germany, the vaccination programmes were started in December 2020. Initially, residents and staff in retirement and care homes were prioritised for vaccination, as outbreaks in these facilities were associated with a particularly high risk of morbidity and mortality (Gharpure, Guo et al. 2021). By March 2021, large parts of this group have been fully vaccinated and the incidence as well as the COVID-19-associated mortality subsequently decreased in this age group.

Despite these positive effects of vaccination, however, there are still groups of people in whom the vaccination response is reduced. These include patients with certain diseases or treatment associated with a reduced immune response, such as renal failure requiring dialysis or immunosuppressive therapy after organ transplantation (Danthu, Hantz et al. 2021, Rincon-Arevalo, Choi et al. 2021, Sattler, Schrezenmeier et al. 2021). Notably, the vaccine response is also reduced in persons of advanced age. Various studies have shown that in persons over 80 years of age, both humoral immunity and cellular immunity are weaker after vaccination against SARS-CoV-2 compared to younger control groups (Collier, Ferreira et al. 2021, Müller, Andrée et al. 2021, Schwarz, Tober-Lau et al. 2021). In a recent study in a home for the elderly, we showed that about 30% of those over 80 years of age showed no neutralisation activity as determined in cell culture three weeks after complete vaccination (Müller, Andrée et al. 2021). Notably, there is growing evidence that a weak humoral immune response associates with an increased infection risk (Feng, Phillips et al. 2021) although other effector mechanisms likely contribute to immune control (Sadarangani, Marchant et al. 2021).

The breadth and magnitude of the immune response has also gained importance in the context of newly emerged virus variants (Harvey, Carabelli et al. 2021). In addition to variants that are primarily associated with an increased risk of transmission (e.g. alpha variant), variants that mediate immune escape are also observed with concern with respect to vaccination (e.g., beta variant). It can be assumed that in the presence of weak or partial immunity, selection and spread of viral variants through the population becomes more relevant. In this context, it is also being discussed whether and at what time or for which group of people a third vaccination will be necessary in order to ensure protection against virus variants (Kamar, Abravanel et al. 2021).

Meanwhile, there are initial reports of outbreaks of SARS-CoV-2 following vaccination in care homes for the elderly (Tober-Lau, Schwarz et al. 2021). The hopeful picture that seems to emerge from this is that the vast majority of infections are mild after complete vaccination. However, it is currently not clear to which extent these infections contribute to boosting immunity against SARS-CoV-2. Here, we describe a large outbreak in a care home for the elderly where almost all residents were fully vaccinated and compare humoral immunity between fully vaccinated individuals and fully vaccinated and infected individuals about 3-4 weeks after infection.

## Patients and Methods

### Patient samples

In early April, the Institute of Virology of the Heinrich-Heine-University was notified by the director of a nursery home for the elderly about a suspected SARS-CoV-2 outbreak after vaccination. In collaboration with local public health authorities, the outbreak was investigated and all residents of the facility where the infections had occurred were offered voluntary blood donations for analysis of anti-SARS-CoV-2 seroprevalence and humoral immune responses. In late April, about 3-4 weeks after the onset of infections, blood samples were collected in accordance with the local public health authority. The retrospective analysis of the outbreak was approved by the ethics committee of the Medical Faculty of the Heinrich-Heine University Duesseldorf, (study no. 2021-1481). When possible, whole-genome sequencing (WGS) of SARS-CoV-2 was performed by the diagnostic laboratory where the initial PCR testing was performed. Characteristics of the study population are summarized in Table 1.

**table 1:** study group.

Tables and legends
| table 1: study group |  |  |  |  |
| --- | --- | --- | --- | --- |
|  | 2 doses | 2 doses + infection | 1 dose + infection | no vaccine + infection |
| studygroup; n | 11 | 13 | 1 | 1 |
| mean age (min/max) | 82.6 (72/90) | 85.3 (68/95) | 81 | 92 |
| gender; n (f/m) | 2/9 | 5/8 | 1/0 | 1/0 |
| Anti-SARS-CoV-2 N positive; n(%) | 11 (84.6%) | 0 (0%) | 1 (100%) | 1 (100%) |
| Anti-SARS-CoV-2 spike IgG; median ( 25/75% percentile) | 268.7 (80.8-394.2) | 6,928 (3,677-40,804) | 766 | 20 |
| Neutralization Titer; median ( 25/75% percentile) | 10 (0-40) | 640 (640-1920) | 40 | 0 |

### Detection of SARS-CoV-2-specific antibodies

Samples were tested for anti-SARS-CoV-2 antibodies using two commercially available test systems. To distinguish antibody responses induced by vaccination from those induced by SARS-CoV-2 infections, the SARS-CoV-2 IgG chemiluminescent microparticle immunoassay (CMIA) from Abbott was performed on an ARCHITECT i2000Abbott. It detects SARS-CoV-2 nucleocapsid-specific (N) IgG antibodies. SARS-CoV-2 spike specific IgG antibodies, induced by either vaccination or infection were measured using the Euroimmun SARS-CoV-2-QuantiVac-ELISA on the Euroimmun Analyzer I-2P according to the manufacturer’s instructions.

A serial dilution endpoint neutralisation test was performed to determine the neutralisation capacity of serum samples against an early SARS-CoV-2 B.1 isolate (GISAID accession ID: EPI_ISL_425126), a SARS-CoV-2 alpha variant (B.1.1.7; GISAID accession ID: EPI_ISL_1209027) and a SARS-CoV-2 beta variant (B1.351; GISAID accession ID: EPI_ISL_1209029). For all isolates, the neutralisation test was performed as previously described (Müller, Andrée et al. 2021, Müller, Ostermann et al. 2021). Briefly, serial dilutions of heat-inactivated (56°C, 30 minutes) serum samples were pre-incubated in cell-free plates with 100 TCID_50_ SARS-CoV2 virus solution for 1 hour at 37° C. After pre-incubation, 100µl of cell suspension containing 7×10^4^/ml Vero cells (ATTC-CCL-81) were added. Plates were incubated at 37°C, 5% CO_2_ for four days before microscopic inspection for virus-induced cytopathic effect (CPE). The neutralisation titre was determined as the highest serum dilution without CPE. Positive, negative, virus only, and cell growth controls were run during each assay.

## Results

In early April, a health care worker with mild symptoms in a care home was tested positive for SARS-CoV-2 by antigen tests. When possible contacts were tested by antigen testing the next day, a second health care worker tested positive, and the remaining tests on eleven individuals were negative. Confirmatory testing of positive antigen test results by PCR was also positive. Both health care workers with SARS-CoV-2 infection were unvaccinated. A few days later, all health care workers and all 42 residents of the care home were tested by antigen tests, of which nine individuals including seven fully vaccinated were positive. During additional testing in the following days a total of 23 residents and four health care workers were tested positive during this outbreak either by antigen test, PCR test or both. Whole viral genome sequences were available from 11 individuals including the two health care workers who were initially tested positive, confirming identical or nearly identical sequences consistent with a single source outbreak with the alpha variant (figure 1). Notably, 18 residents were fully vaccinated and showed no or only mild symptoms. Four residents were unvaccinated including one with a severe disease course who later deceased in the hospital. One resident had only received the first vaccine dose.

**Figure 1:**
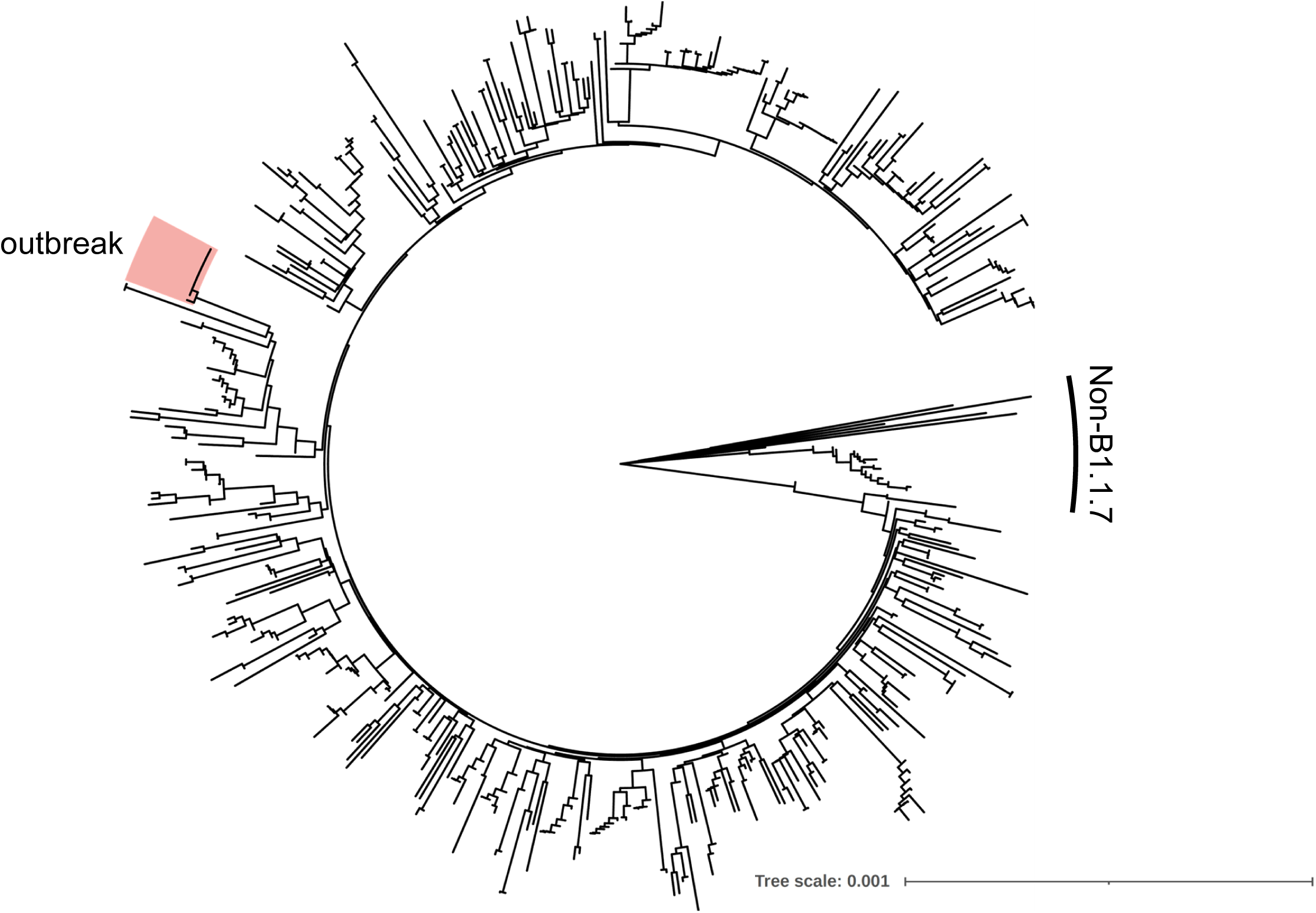
Phylogenetic tree of the outbreak sequences with local SASR-CoV2 isolates. Whole SARS-CoV-2 genome sequences from the outbreak were aligned with 506 local SARS-CoV-2 isolates sampled during the same week (provided by (Walker, Houwaart et al. 2021)) and a phylogenetic tree was calculated by the Neighbor-Joining method using the Tamura-Nei genetic distance model utilizing the software Geneious R.10.2.6 and visulized with iTOL6.3 (Letunic and Bork 2016). The outbreak samples are highlighted in red. Non-B1.1.7 sequences are indicated.

Three to four weeks later, blood samples were collected from 26 residents for analysis of the humoral immune response including 15 residents with a documented infection by PCR and/or antigen testing and 11 residents who did not test positive. While all 11 residents previously tested negative by PCR and antigen testing had no detectable anti-SARS-CoV-2 nucleocapsid antibodies, 13 of the 15 residents previously tested positive had detectable anti-SARS-CoV-2 nucleocapsid antibodies. Collectively, the serology results were consistent with the PCR and antigen test results with only two exceptions. Of note, both residents with undetectable anti-SARS-CoV-2 nucleocapsid antibodies were tested positive by PCR. Of the 26 residents, 24 were fully vaccinated at least eight weeks before sampling and at least five weeks before exposure. One individual refused vaccination and one additional individual received only one vaccine dose. The mean age of the fully vaccinated and infected group was 85.3 years and of the uninfected group 82.6 years (table 1).

The anti-SARS-CoV-2 spike response was determined by ELISA quantifying binding antibody units (BAU/ml). The median anti-SARS-CoV-2 spike antibody levels in the vaccinated and uninfected group was 268.7 BAU/ml including one outlier with a substantially higher level (16,862 BAU/ml). Importantly, the median anti-SARS-CoV-2 spike level was significantly higher in the fully vaccinated and infected group (6,928 BAU/ml). The anti-SARS-CoV-2 spike levels were on average 6.6-fold higher after infection than in the uninfected group (figure 2A). Notably, the individual who had received only a single vaccine dose had intermediate level of 766 BAU/ml an and the unvaccinated but infected individual had a very low anti-SARS-CoV-2 spike level (20 BAU/ml).

**Figure 2:**
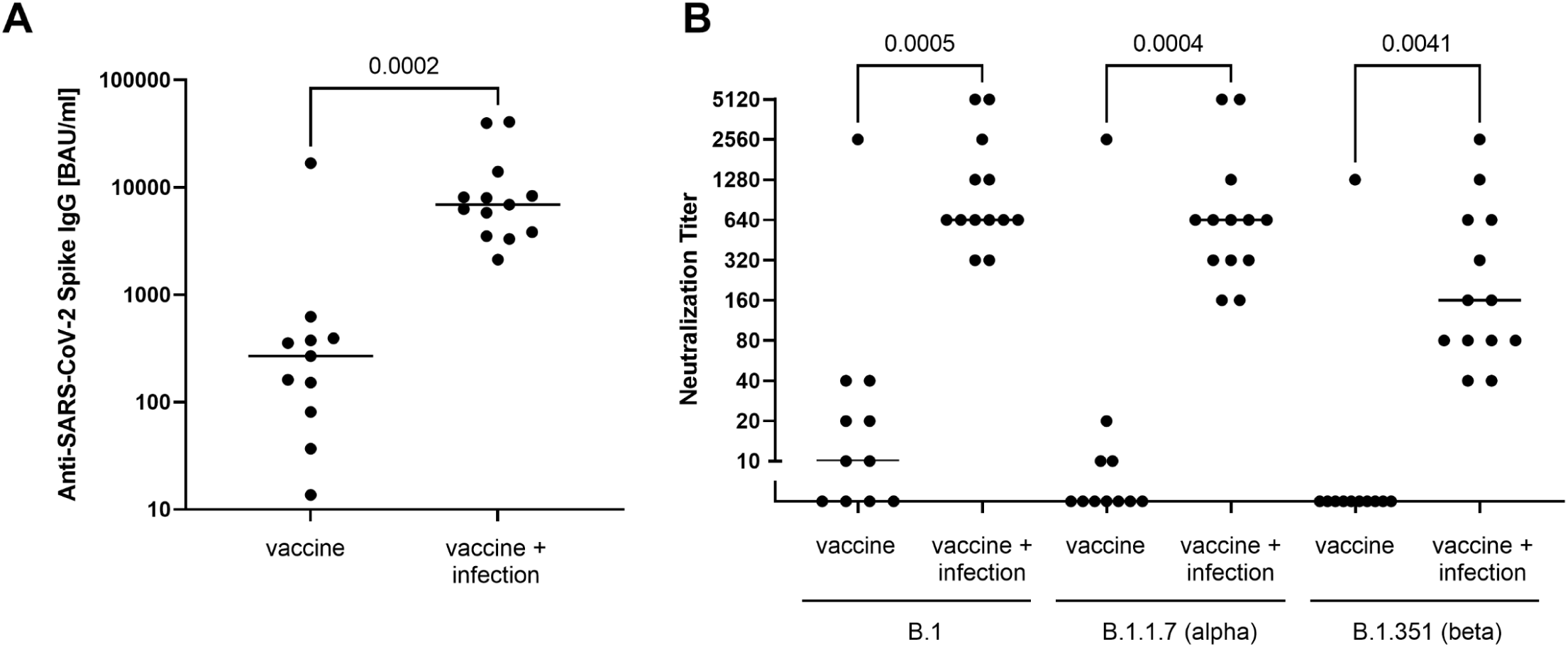
Humoral immune response against SARS-CoV-2. (A) anti-SARS-CoV-2 spike IgG titres were measured as binding antibody units/ml (BAU/ml) using Euroimmun anti-SARS-CoV-2-QuantiVac-ELISA. For comparison of the two groups, a Mann-Whitney test was performed with the p-value depicted in the figure. (B) To determine the anti-SARS-CoV-2 neutralisation capacity against SARS-CoV-2 B.1, SARS-CoV-2 B.1.1.7 (alpha variant), SARS-CoV-2 B.1.351 (beta variant), a serial dilution endpoint neutralisation test was performed for the two groups. The reciprocal of the NT titre is depicted, bars represent the respective median. For comparison of the groups, a one-way ANOVA (Kruskal-Wallis test) with Dunn’s correction for multiple comparisons was performed. The p-values are depicted in the figure.

The results of the ELISA were confirmed in a neutralisation assay using a replication competent SARS-CoV-2 B.1-isolate in cell culture. While all fully vaccinated and infected individuals showed high neutralisation titres the mean titre was significantly lower in the uninfected but fully vaccinated group, including four of eleven individuals with no neutralizing activity (figure 2B). When the activity against the alpha and beta variant was determined, neutralisation declined and the frequency of individuals lacking neutralisation activity further increased (7 of 11 alpha variant, 10 of 11 beta variant), consistent with immune escape of the beta variant. Notably, although there was also evidence for decreased neutralisation titres against the beta variant, the group of fully vaccinated and infected individuals all showed high neutralisation capacity against all three variants tested here. Collectively, the serological results suggest a strong booster effect of SARS-CoV-2 infection in fully vaccinated individuals consistent with a protective effect also against immune escape variants.

## Discussion

SARS-CoV-2 infections after vaccination have been described in various reports and demonstrate that sustained sterile immunity against COVID-19 is not always achieved. The outbreak described here in a nursing home for the elderly described here demonstrates that a super-spreading event can also occur even in the context of full vaccination. Given the short duration of the outbreak event, a reliable reconstruction of the infection chain is challenging. The first cases of infection had been detected in two non-vaccinated staff members of the facility when they had developed symptoms. A possible common source of infection for these two cases could not be found. The further timing of infections among facility residents suggests that at least one of the unvaccinated staff members and possibly both were responsible for transmission to secondary cases. Nearly all secondary cases of infection had contact with these two staff members in the phase prior to initial diagnosis. Phylogenetic analysis of the available complete viral genomes confirmed the assumption of an outbreak in which the isolates from staff and residents are almost identical and form a distinct cluster to all regionally circulating alpha variants studied. During the outbreak, a total of 27 infections could be detected by PCR and/or antigen test within two weeks, 19 of them in fully vaccinated persons. Although it cannot be formally ruled out with certainty, there was no evidence from the chronology that the secondary cases of infection had originated from vaccinated individuals.

The outbreak was caused by the alpha variant (B.1.1.7), which was the dominant variant in the region at that time, accounting for about 90% of the infection cases. In the outbreak isolate, an additional substitution in the spike gene was detected, which leads to an amino acid exchange at position 779 (glutamine to lysine). Based on the localisation outside the receptor binding domain in the S2 domain of the spike protein, we speculate that this substitution has no strong influence on the efficacy of humoral immunity. This substitution has also not been associated with immune escape in the context of known variants of concern. In publicly available databases, this substitution has been described only in single isolates and is not present in 4095 isolates sequenced from regional infections (Walker, Houwaart et al. 2021). Although experimental confirmation is pending, we do not propose that the Q779K substitution promoted the outbreak after full vaccination.

The size of the outbreak allowed us to comparatively analyse the antibody response of vaccinated individuals with and without infection. On one hand, the detection of antibodies against the nucleocapsid confirmed contact with the virus in an indirect detection method in 13 of 15 infections. At the same time, in all 11 cases in which there was no evidence for infection either anamnestically or in direct virus detection assays, the test for nucleocapsid-specific antibodies was negative. Testing of nucleocapsid-specific antibodies in vaccinated persons, therefore, may provide additional information whether an infection has occurred, although the absence of these antibodies does not reliably rule out an infection. There were considerable differences in the quantitative comparison of the antibodies against the spike protein. In all cases with infection despite vaccination, extremely high antibody levels and a very high neutralisation activity were measured. In contrast, antibody levels and neutralisation titres were significantly lower in the vaccinated individuals without infection, with one exception. This person had high antibody levels comparable to the vaccinees after infection. Although multiple antigen tests and PCR tests were negative, it cannot be excluded that an infection was missed in this case.

The high antibody levels most likely reflect a boost to the immune system by the infection. Unfortunately, samples from before the infection are not available, so it is not possible to exactly determine the extent of immune boosting by the infection. It is also not clear whether immunity was weaker in the group of infected people before infection than in the group without infection. Even among those individuals without infection, there were contacts with the suspected superspreaders, so exposure seems possible. The lack of longitudinal data as well as a control group of persons without vaccination examined in the same period after infection are important limitations of this study. It is noteworthy that the neutralisation titres were on average significantly lower in unvaccinated convalescents analysed at our lab with the same assay compared to vaccinated and infected individuals (data not shown).

Even after complete vaccination, neutralising antibodies are not detectable in all individuals. Especially in elderly and immunosuppressed people, immunity after vaccination is less pronounced. In a previous study, about 30% of those over 80 years of age had no neutralising activity against the prototype virus three weeks after the second dose of an mRNA vaccine (Müller, Andrée et al. 2021). In this study, the antibody response was at the same low level in vaccinated individuals at comparable ages about eight weeks after vaccination. Infection after vaccination, as in this case with the alpha variant of SARS-CoV-2, thus appears to strongly boost immunity. It should be emphasised that vaccinated individuals showed considerable cross-reactivity after infection, which also neutralises the beta variant. The functional immune escape of the beta variant is particularly evident in the loss of neutralisation activity at low antibody levels. It is therefore reasonable to assume that boosting of immunity by an infection may provide additional protection against variants. Although it seems possible that the same effect could be achieved by a third vaccine dose, the observation also has implications for a situation in which SARS-CoV-2 becomes endemic and contact with the virus occurs regularly. In addition, infection may confer sterilizing immunity in the respiratory tract in vaccinees which is less likely to be achieved with currently licensed vaccines in Germany. A prerequisite for the acceptance of infections would be a safe protection against complications and severe disease courses. Even though data are not yet sufficient, severe courses of COVID-19 in fully vaccinated individuals seem to be rare so far (Haas, Angulo et al. 2021, Hall, Foulkes et al. 2021).

It should be highlighted that only the humoral immune response was investigated in this study. The relevance of cellular immunity by T cells was not part of the analysis and is certainly also of great importance. This retrospective study also does not allow any conclusions about the causal mechanisms that led to the infections after vaccination. This would require longitudinal studies in vaccinated persons up to the time of a subsequent infection. In summary, the results confirm the observation that SARS-CoV-2 outbreaks are possible in a fully vaccinated risk group, but these are associated with rather low disease severity. Furthermore, we conclude that humoral immunity is significantly enhanced after infection in a vaccinated risk group and also protects against immune escape variants, at least in vitro.

## Data Availability

Not applicable

## Acknowledgments

We thank Yvonne Dickschen for excellent technical assistant. This work was supported by the VIRus ALliance NRW (VIRAL) from the Ministry of Culture and Science of the State of North Rhine Westphalia (323-8.03-151826) and the Jürgen Manchot Foundation.

